# Racial, Economic and Health Inequality and COVID-19 Infection in the United States

**DOI:** 10.1101/2020.04.26.20079756

**Authors:** Vida Abedi, Oluwaseyi Olulana, Venkatesh Avula, Durgesh Chaudhary, Ayesha Khan, Shima Shahjouei, Jiang Li, Ramin Zand

## Abstract

**Background:** There is preliminary evidence of racial and social-economic disparities in the population infected by and dying from COVID-19. The goal of this study is to report the associations of COVID-19 with respect to race, health and economic inequality in the United States.

**Methods:** We performed a cross-sectional study of the associations between infection and mortality rate of COVID-19 and demographic, socioeconomic and mobility variables from 369 counties (total population: 102,178,117 [median: 73,447, IQR: 30,761-256,098]) from the seven most affected states (Michigan, New York, New Jersey, Pennsylvania, California, Louisiana, Massachusetts).

**Findings:** The risk factors for infection and mortality are different. Our analysis shows that counties with more diverse demographics, higher population, education, income levels, and lower disability rates were at a higher risk of COVID-19 infection. However, counties with higher disability and poverty rates had a higher death rate. African Americans were more vulnerable to COVID-19 than other ethnic groups (1,981 African American infected cases versus 658 Whites per million). Data on mobility changes corroborate the impact of social distancing.

**Interpretation:** The observed inequality might be due to the workforce of essential services, poverty, and access to care. Counties in more urban areas are probably better equipped at providing care. The lower rate of infection, but a higher death rate in counties with higher poverty and disability could be due to lower levels of mobility, but a higher rate of comorbidities and health care access.

## INTRODUCTION

The complexity of managing patients with the Coronavirus disease 2019 (COVID-19), a global pandemic^1^ originated in China,^2^ has led to the widespread implementation of preventative measures such as social distancing and mask-use^3^ in many countries including the United States (US). As of April 14, 2020, there were over 1.9 million confirmed cases around the world with 601,000 cases and 24,129 deaths in the US alone.^4^ It has been reported that age 65 and older, body mass index ≥ 40, immunosuppression, smoking, hypertension, cardiovascular diseases are underlying conditions that increase the risk of death from COVID-19.^5,6^

The most recent conundrum of this disease is ascribed to the preliminary evidence of racial disparities in the population infected and dying from COVID-19.^3^ In a recent study, the center for disease control and prevention (CDC) in the US reported data from fourteen states and suggested that the US Black population may be disproportionally affected by COVID-19.^3^ This observation is consistent with the influenza A (H1N1) pandemic where other studies showed evidence of racial and ethnic disparities in the population affected both in exposure, severity, and mortality of the disease.^7,8^ As states release the racial and ethnic demographic data of COVID-19 cases, in addition to the increased spread of this disease to the central states, it is imperative that we understand the patterns of infection and death to reduce the risks, especially for high-risk population, and resolve issues that impede the provision of optimal care.

In this study, we conducted a population-based analysis to explore racial and economic inequality associated with the infection rate and risk of mortality due to COVID-19 in the US. The goal of the study is to provide evidence on the association of COVID-19 with respect to race, income level, poverty, education, and the impact of preventative measures such as social distancing. We trust that the decision making by the states’ officials will be driven by data and based on their unique needs and population characteristics to help in combating this disease.

## METHODOLOGY

The study was conducted at two levels: 1) Analysis of population characteristics (44 variables) for 369 counties in seven states which had the highest rate of COVID-19 infection as of April 9^th^, 2020 along with COVID-19 infection and mortality rates. The included states were California, Michigan, New York, New Jersey, Louisiana, Pennsylvania, and Massachusetts; 2) Analysis of COVID-19 related infection and death rate across all the states in the US with race/ethnicity information on the affected subject when available.

### Data source

Data sources in this study include, 1) publicly available data from USAfacts and the US Census Bureau for COVID-19 cases and county-level demographic data,^9,10^ 2) COVID-19 data reported by each state on their department of health websites,^9^ 3) State Population by Race/Ethnicity data,^11,12^ and 4) mobility data extracted from Google.^13^

Variables used in this study include county-level information on total population, mobility, race, poverty level, median income, education, disability, rate of the insured population. Mobility data were extracted from Google as reported on April 05, 2020. The state-level data were extracted on April 16, 2020. The outcome variables include the rate of COVID-19 infection and all COVID-19 related death as provided by each state’s department of health as of April 09, 2020. The infection rate is based on the reported results from all the laboratories testing samples in each county/state. The mortality data are reported by hospitals, nursing homes, and other health facilities. Table 1 summarizes the data elements used in this study. Only data provided by the states on their official websites were included in this study. Additionally, to compare the rate of COVID-19 cases and death, the population data for each ethnic/racial group affected were extracted from health department websites.

**Table 1.**
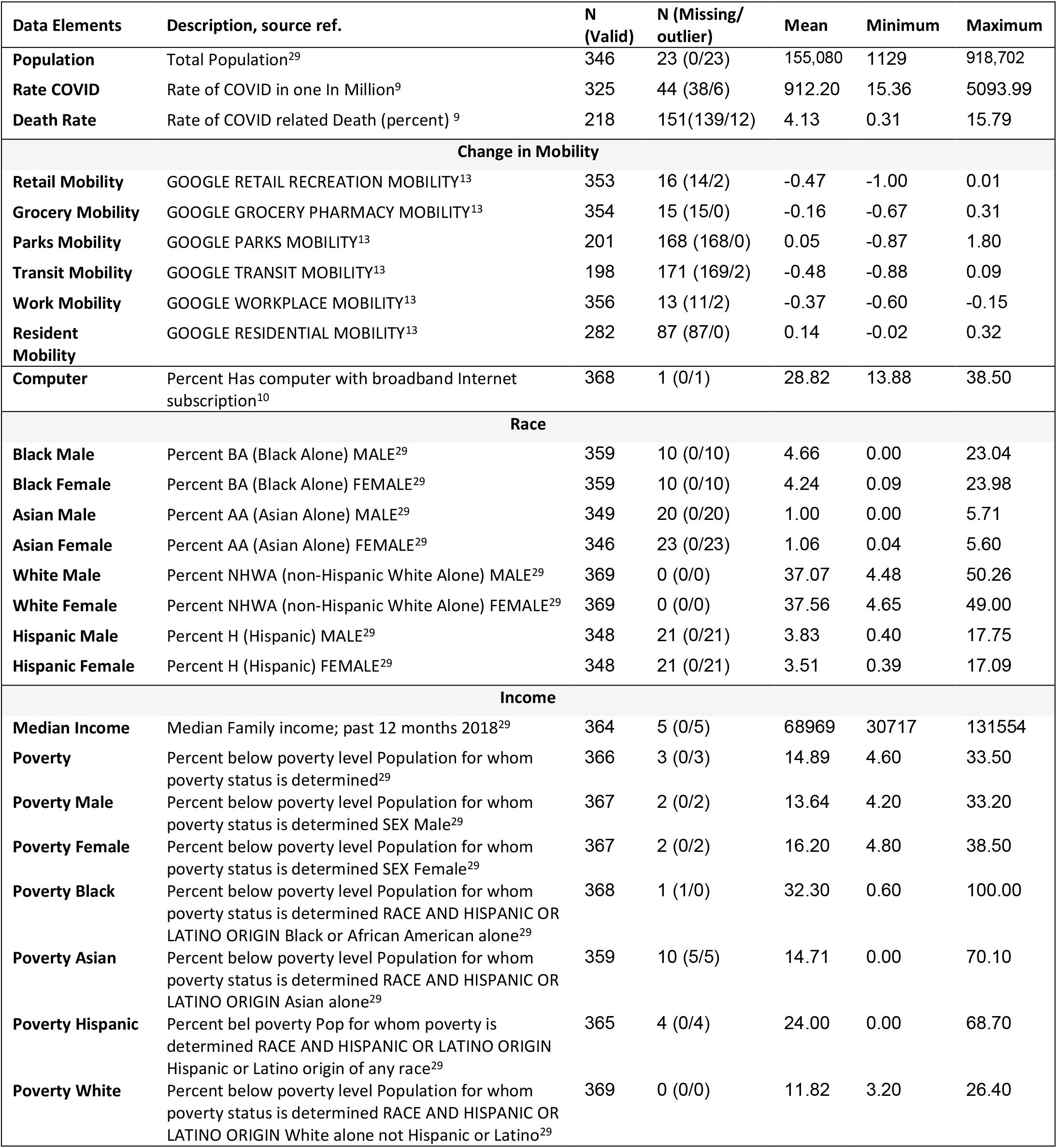
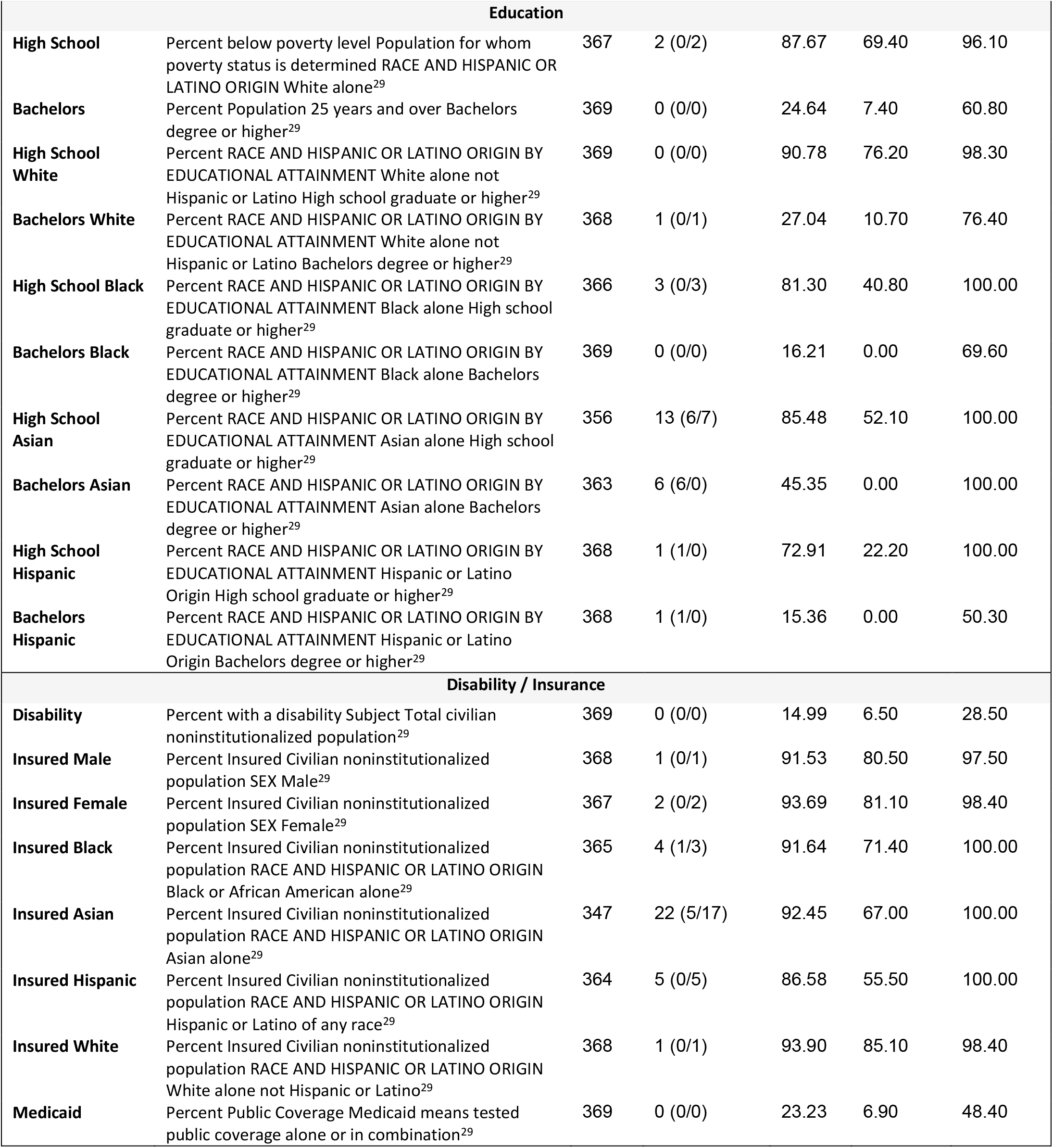
Data elements, descriptions, reference to the source of data, and base statistics used in this study.

### Statistical analysis

We summarized all continuous variables as mean ± standard deviation or median with inter-quartile range [IQR], and categorical variables as percentages. Data from different sources were extracted and analyzed for outliers. Values not within three inter-quartiles were removed as part of the data pre-processing. Each continuous variable was centralized, and z-score transformed. Thus, the transformed variables passed the normality test and the correlation matrix was created. Bivariate, partial correlation, and regression were used to test hypotheses of association. The correlation coefficients between “Death Rate” or “Infection Rate” with independent variables were calculated by Pearson’s correlation (R corr package). Partial correlation was further evaluated by Pearson’s correlation (R ppcor package) to determine if the existing correlation was still valid after controlling the second independent variable. Bonferroni correction for multiple testing of controlling variables was considered to adjust the p-value of the correlation. Bivariate linear regression adjusted for “State” variables was utilized to test the association between “Death Rate” or “Infection Rate” with independent variables. The raw p-value was present in the forest plot. False discovery rate (FDR) corrections for multiple testing were calculated using the Benjamin and Hochberg procedure. Statistical analyses were performed using R version 3.6.2.^14^ and IBM SPSS Statistics 26.^15^

## RESULTS

### Population, mobility, and socioeconomic determinants

We extracted data from four different sources on 369 counties from seven states, including five states from the East Coast (Michigan, New York, New Jersey, Pennsylvania, and Massachusetts), one state from the West Coast (California), and one state from the South (Louisiana) with the total population of 102,178,117 (median: 73,447, IQR: 30,761-256,098). The information on race, income, education level, insurance, poverty, and disability including the description of abbreviated variables are summarized in Table 1, (Supplemental Table S1 includes additional summary statistics of the dataset).

Our data show a significant association among different socioeconomic determinants, such as poverty level, education, and income (See Table S2). In particular, counties with a higher percentage of people below the poverty level had a significantly lower percentage of the population with higher education (Pearson correlation: −0.52, p<0.005 for Bachelor’s degree; Pearson correlation: −0.61, p<0.005 for High School), as well as a lower percentage of people insured, but a higher percentage of people on Medicaid (Pearson correlation: 0.77, p<0.005) or on disability (Pearson correlation: 0.41, p<0.005; See Table S2 for more details). Counties with a higher percentage of residents below the poverty level had a higher percentage of Blacks (Pearson correlation: 0.52, p<0.005 for men; Pearson correlation: 0.50, p<0.005 for women) and a lower percentage of non-Hispanic Whites (Pearson correlation: −0.30, p<0.005 for men; Pearson correlation: −0.33, p<0.005 for women).

### Counties with a higher total population, more diverse demographics, higher education, and income level are at a higher risk of COVID-19 infection

The COVID-19 infection rate per one Million (mean: 912.20±1,034.26) ranged from 15.36 to 5,093.99 in different counties (Table 1 and S1). Figure 1 shows the map of Pennsylvania with total population for each county, rate of infection and death due to COVID-19 infection, as well as, median income in the counties and percentage of the population who are identified as non-Hispanic Whites. The map of the other six states is provided as Supplemental Figure S1-S6 for reference. The outliers were not removed in these figures.

**Figure 1.**
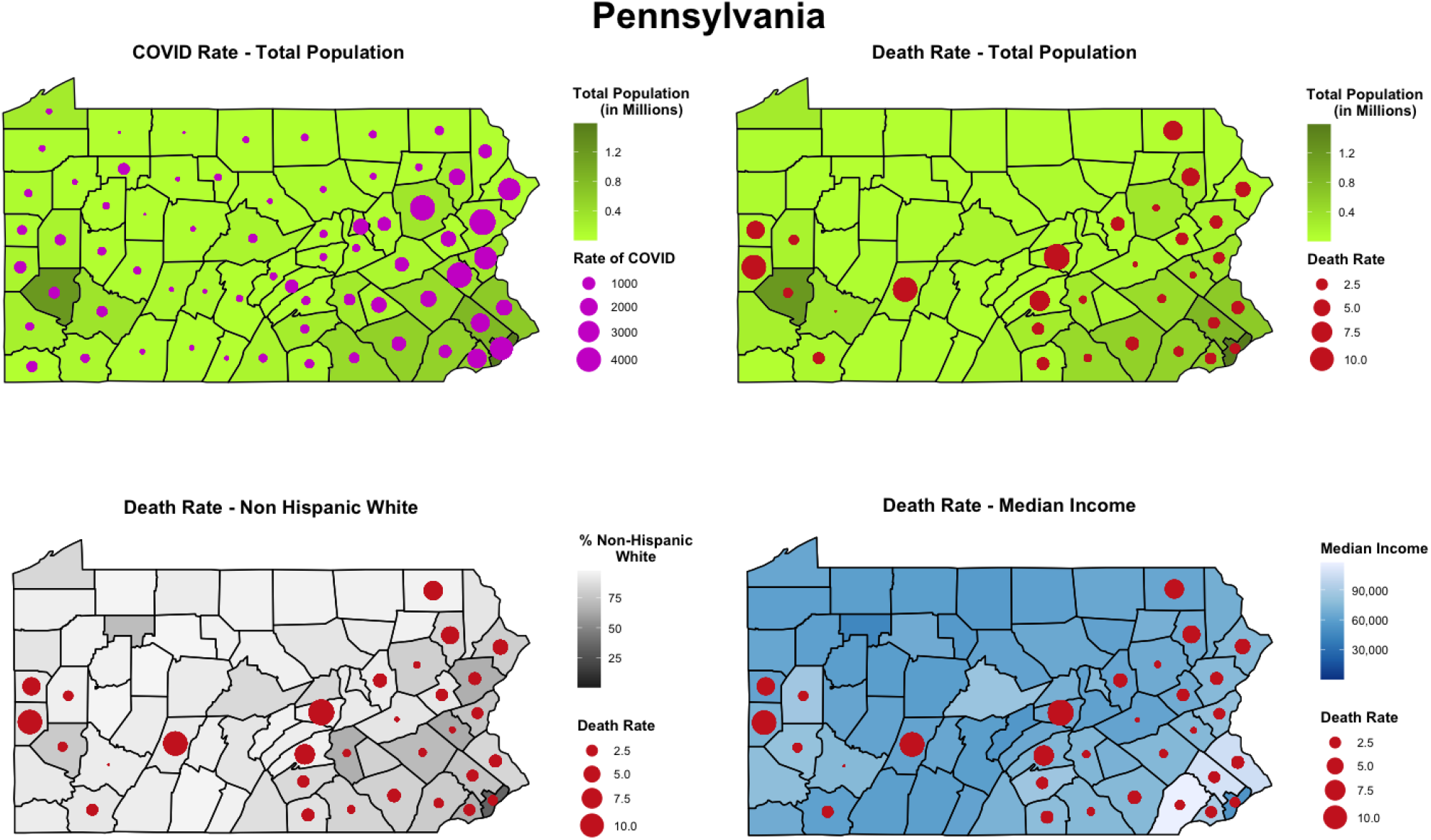
Population count, non-Hispanic White, and Median Income and the rate of COVID-19 and related death in the counties of the state of Pennsylvania, as of April 9^th^, 2020.

The results of the bivariate linear regression (Figure 2, Table S3) estimate effect sizes (regression coefficients) of a number of variables contributed to COVID-19 infection when controlled for states in the model. Counties with a higher population (est. 0.34, 95% CI: 0.24, 0.44, q<1.1E-08), a higher median income (est. 0.36, 95% CI: 0.25, 0.48, q<2.3E-08) as well as a more diverse population (higher percentage of Hispanics, Asians, and Blacks) have a higher rate of infection. More specifically, a higher percentage of Asians (est. 0.32, 95% CI: 0.20, 0.44 q<6.3E-07, women; est. 0.32, 95% CI: 0.20, 0.43, q<4.5E-07, men), Blacks (est. 0.47, 95% CI: 0.32, 0.62, q<2.3E-08, women; est. 0.35, 95% CI: 0.20, 0.51, q<1.7E-05, men), as well as Hispanics (est. 0.49, 95% CI: 0.34, 0.64, q<1.2E-08, women; est. 0.46, CI: 0.31, 0.62,, q<8.1E-08, men) are associated with a higher rate of infection while a higher percentage of non-Hispanic Whites (est. −0.41, 95% CI: - 0.55, −0.26, q<2.9E-07, women; est. −0.44, 95% CI: −0.58, −0.30, q<4.2E-08, men) is associated with a lower rate of COVID-19. Change in grocery mobility (est. −0.24, 95% CI: −0.36, −0.13, q<5.5E-05), retail mobility (est. −0.26, 95% CI: −0.38, −0.14, q<4.5E-05) as well as work mobility (est. −0.31, 95% CI: −0.43, −0.20, q< 9.0E-07) were associated with lower rate of infection. Another protective factor in terms of rate of infection for the counties analyzed was a higher percentage of disability (est. −0.159, 95% CI: −0.265, −0.053, q<0.006). We also analyzed the rate of infection for counties with respect to their percentage of uninsured and found no significant association other than among men (est. 0.181, 95% CI: 0.05, 0.313, q<1.2E-02) and non-Hispanic Whites (est. 0.251, 95% CI: 0.123, 0.380, q<3.1E-04). Furthermore, in our stepwise regression model, minorities specifically Black and Hispanic women, poverty, and level of education among non-Hispanic Whites, disability, the total county population, and level of mobility are predictors of the rate of COVID-19 infection (Table S4).

**Figure 2.**
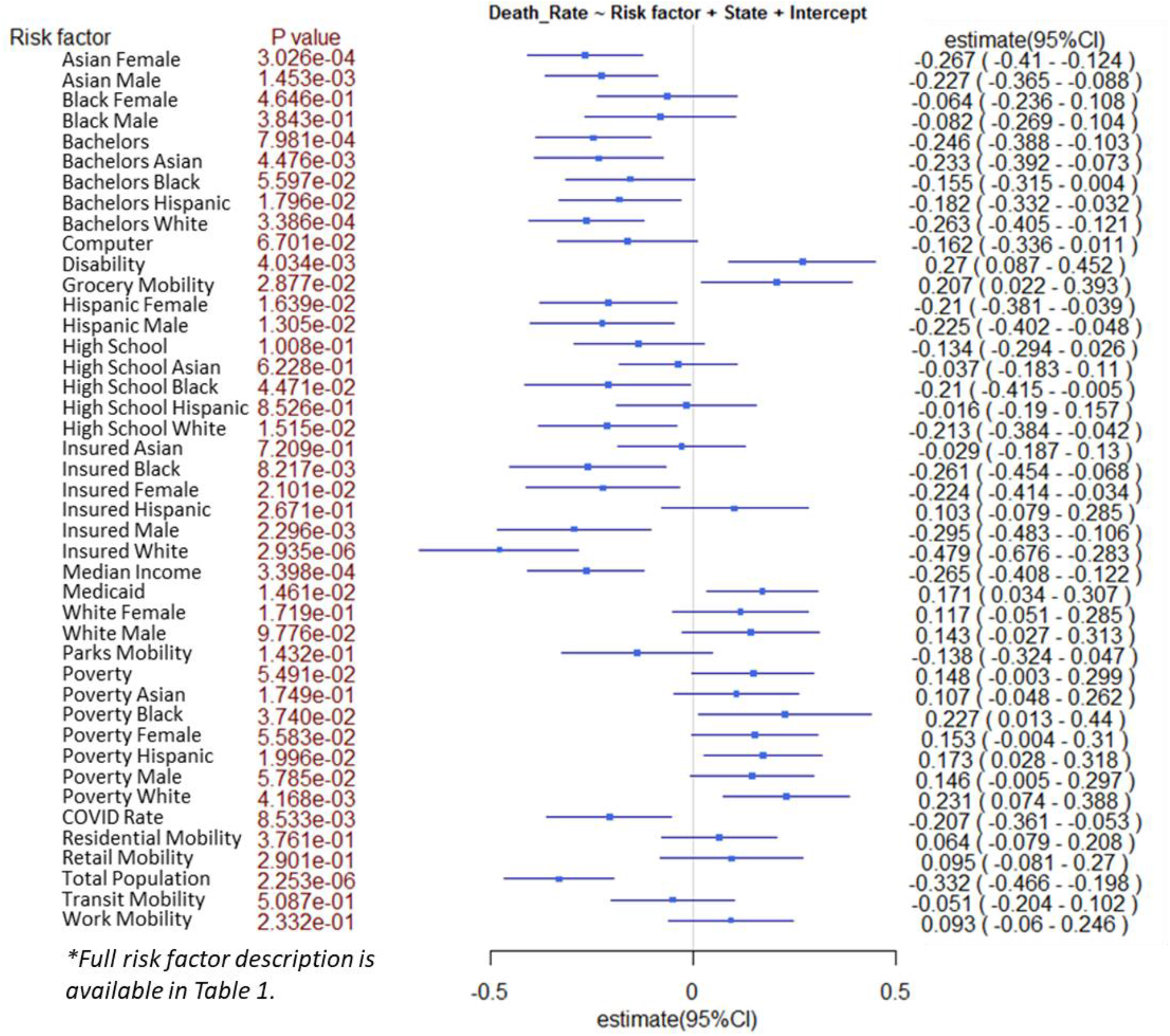
Bivariate analysis of risk factors for infection by COVID-19.

### Counties with a smaller population, higher poverty levels, and higher disability have a higher rate of mortality

The COVID-19 related death (mean: 4.13% ± 2.70%, median: 3.40, IQR: 2.22-5.61) varied among different counties (Table 1 and S1). Figure 3 and Table S5 show the results of the bivariate regression analysis estimating the odds of mortality due to COVID-19 infection (model corrected for states). Protective factors for the counties are a higher percentage of Asians (est. −0.27, 95% CI: −0.41, −0.12, q< 0.003, women; est. −0.23, 95% CI: −0.37, −0.09, q<0.009, men) and education level with a bachelor’s degree or higher with an odds ratio ranging from −0.41 to −0.03 across the various ethnicities (see Figure 3). Other protective factors for counties include having a higher percentage of people insured (strongest indicator being for non-Hispanic White people with an estimate of −0.48, 95% CI: −0.68, −0.28, q<6.0E-05), as well as median income (est. −0.27, 95% CI: −0.41, −0.12, q<0.003). The total population in the counties is also a major indicator (est. −0.33, 95% CI: −0.43, −0.20, q<6.0E-05) of lower COVID related death. We have also explored the association between total population and various confounding factors, such as mobility data when analyzing the death rate and found that the total population is still an important protective factor (see Table S6, Figure S7 and S8). Factors significantly associated with higher mortality in the counties analyzed include a higher percentage of people under the poverty level (for all the races analyzed in this study), a higher percentage of people on Medicaid (est. 0.17, 95% CI: 0.03, 0.30, q<0.04) as well as a higher rate of people with disability in the county (est. 0.27, 95% CI: 0.09, 0.45, q<0.02). Grocery mobility was also highly associated with mortality (est. 0.21, 95% CI: 0.02, 0.39, q<0.06).

**Figure 3.**
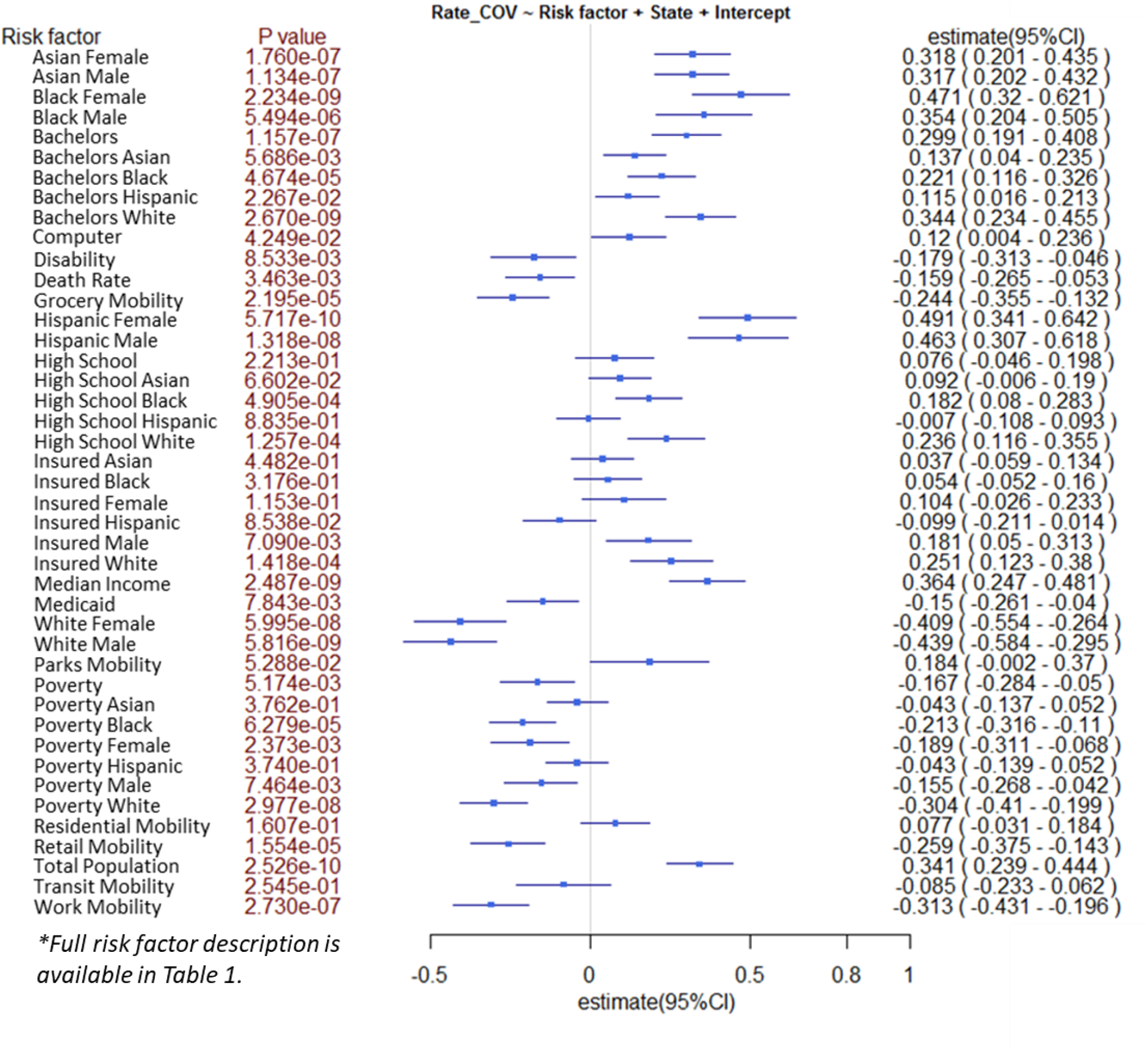
Bivariate analysis of risk factors for mortality due to COVID-19.

To better understand the characteristics of counties with higher or lower death rates, we performed a comparative analysis using ANOVA and found that, similar to the above, counties with more population diversity, higher income and education, a lower rate of disability, and a higher rate of the insured population have a significantly lower than the median death rate. Table 2 and Table S7 summarizes the population characteristics when counties are compared with death rate lower and higher than the median (median death rate is 3.4% across the counties in the seven states, Table S1). Park and retail mobility changes are significantly different between the two groups. The average number of Asians (both man and woman), as well as Hispanics (both man and woman), is significantly higher (p<0.05) in group 1 (Death Rate ≤ 3.4). The counties with higher death rates have lower median income and higher poverty levels across all the races. The group with a lower death rate has also a higher rate of the insured population and a lower rate of disability. The percentage of Medicaid is significantly higher in the group with a higher death rate.

**Table 2.**
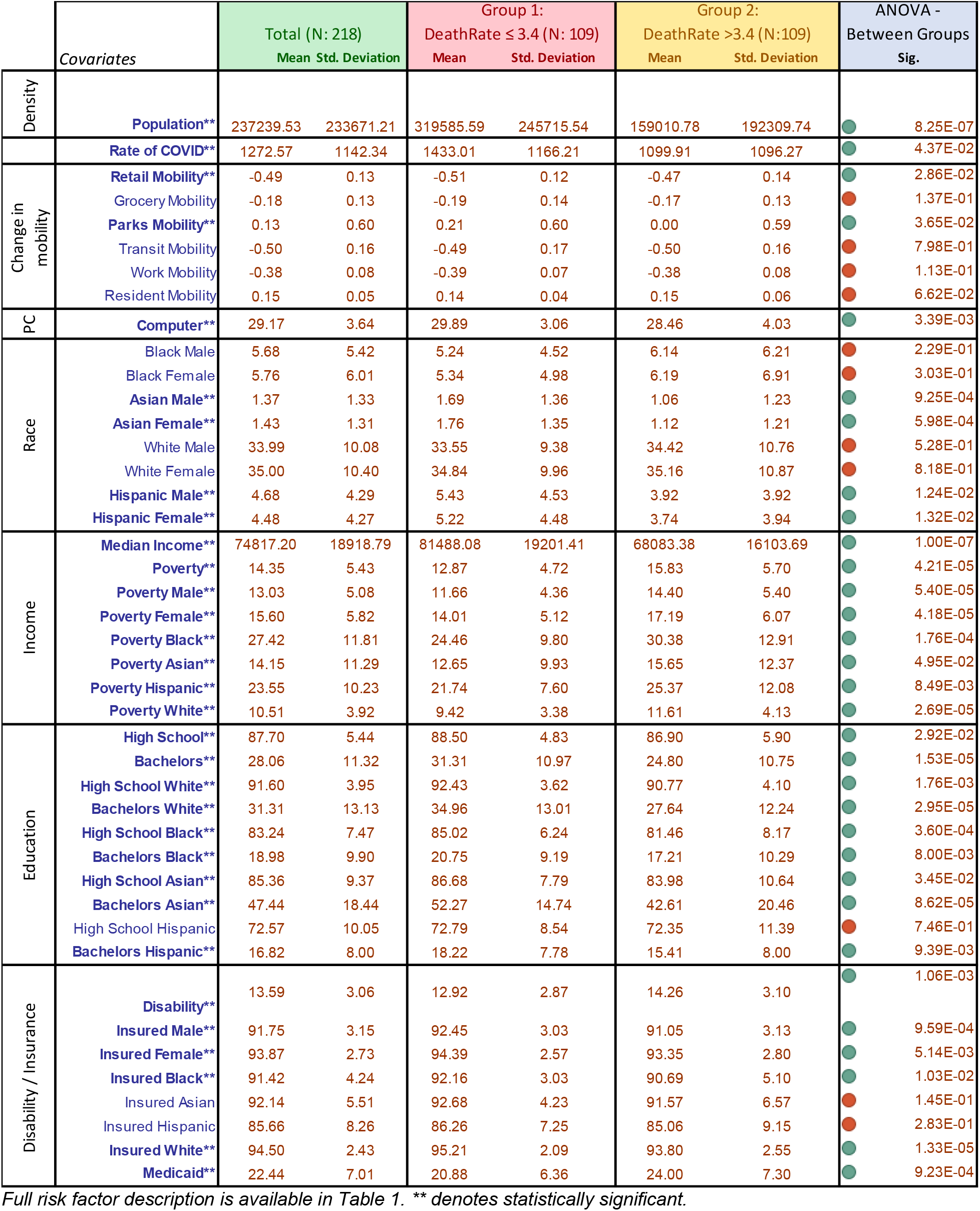
ANOVA comparing counties with higher and lower than the median death rate.

|  | Covariates | Total (N: 218)<br>Mean Std. Deviation |  | Group 1:<br>DeathRate ≤ 3.4 (N: 109)<br>Mean Std. Deviation |  | Group 2:<br>DeathRate >3.4 (N:109)<br>Mean Std. Deviation |  | ANOVA -<br>Between Groups<br>Sig. |  |
| --- | --- | --- | --- | --- | --- | --- | --- | --- | --- |
| Density | Population** | 237239.53 | 233671.21 | 319585.59 | 245715.54 | 159010.78 | 192309.74 |  | 8.25E-07 |
|  | Rate of COVID** | 1272.57 | 1142.34 | 1433.01 | 1166.21 | 1099.91 | 1096.27 |  | 4.37E-02 |
| Change in mobility | Retail Mobility** | -0.49 | 0.13 | -0.51 | 0.12 | -0.47 | 0.14 |  | 2.86E-02 |
|  | Grocery Mobility | -0.18 | 0.13 | -0.19 | 0.14 | -0.17 | 0.13 |  | 1.37E-01 |
|  | Parks Mobility** | 0.13 | 0.60 | 0.21 | 0.60 | 0.00 | 0.59 |  | 3.65E-02 |
|  | Transit Mobility | -0.50 | 0.16 | -0.49 | 0.17 | -0.50 | 0.16 |  | 7.98E-01 |
|  | Work Mobility | -0.38 | 0.08 | -0.39 | 0.07 | -0.38 | 0.08 |  | 1.13E-01 |
|  | Resident Mobility | 0.15 | 0.05 | 0.14 | 0.04 | 0.15 | 0.06 |  | 6.62E-02 |
| PC | Computer** | 29.17 | 3.64 | 29.89 | 3.06 | 28.46 | 4.03 |  | 3.39E-03 |
| Race | Black Male | 5.68 | 5.42 | 5.24 | 4.52 | 6.14 | 6.21 |  | 2.29E-01 |
|  | Black Female | 5.76 | 6.01 | 5.34 | 4.98 | 6.19 | 6.91 |  | 3.03E-01 |
|  | Asian Male** | 1.37 | 1.33 | 1.69 | 1.36 | 1.06 | 1.23 |  | 9.25E-04 |
|  | Asian Female** | 1.43 | 1.31 | 1.76 | 1.35 | 1.12 | 1.21 |  | 5.98E-04 |
|  | White Male | 33.99 | 10.08 | 33.55 | 9.38 | 34.42 | 10.76 |  | 5.28E-01 |
|  | White Female | 35.00 | 10.40 | 34.84 | 9.96 | 35.16 | 10.87 |  | 8.18E-01 |
|  | Hispanic Male** | 4.68 | 4.29 | 5.43 | 4.53 | 3.92 | 3.92 |  | 1.24E-02 |
|  | Hispanic Female** | 4.48 | 4.27 | 5.22 | 4.48 | 3.74 | 3.94 |  | 1.32E-02 |
| Income | Median Income** | 74817.20 | 18918.79 | 81488.08 | 19201.41 | 68083.38 | 16103.69 |  | 1.00E-07 |
|  | Poverty** | 14.35 | 5.43 | 12.87 | 4.72 | 15.83 | 5.70 |  | 4.21E-05 |
|  | Poverty Male** | 13.03 | 5.08 | 11.66 | 4.36 | 14.40 | 5.40 |  | 5.40E-05 |
|  | Poverty Female** | 15.60 | 5.82 | 14.01 | 5.12 | 17.19 | 6.07 |  | 4.18E-05 |
|  | Poverty Black** | 27.42 | 11.81 | 24.46 | 9.80 | 30.38 | 12.91 |  | 1.76E-04 |
|  | Poverty Asian** | 14.15 | 11.29 | 12.65 | 9.93 | 15.65 | 12.37 |  | 4.95E-02 |
|  | Poverty Hispanic** | 23.55 | 10.23 | 21.74 | 7.60 | 25.37 | 12.08 |  | 8.49E-03 |
|  | Poverty White** | 10.51 | 3.92 | 9.42 | 3.38 | 11.61 | 4.13 |  | 2.69E-05 |
| Education | High School** | 87.70 | 5.44 | 88.50 | 4.83 | 86.90 | 5.90 |  | 2.92E-02 |
|  | Bachelors** | 28.06 | 11.32 | 31.31 | 10.97 | 24.80 | 10.75 |  | 1.53E-05 |
|  | High School White** | 91.60 | 3.95 | 92.43 | 3.62 | 90.77 | 4.10 |  | 1.76E-03 |
|  | Bachelors White** | 31.31 | 13.13 | 34.96 | 13.01 | 27.64 | 12.24 |  | 2.95E-05 |
|  | High School Black** | 83.24 | 7.47 | 85.02 | 6.24 | 81.46 | 8.17 |  | 3.60E-04 |
|  | Bachelors Black** | 18.98 | 9.90 | 20.75 | 9.19 | 17.21 | 10.29 |  | 8.00E-03 |
|  | High School Asian** | 85.36 | 9.37 | 86.68 | 7.79 | 83.98 | 10.64 |  | 3.45E-02 |
|  | Bachelors Asian** | 47.44 | 18.44 | 52.27 | 14.74 | 42.61 | 20.46 |  | 8.62E-05 |
|  | High School Hispanic | 72.57 | 10.05 | 72.79 | 8.54 | 72.35 | 11.39 |  | 7.46E-01 |
|  | Bachelors Hispanic** | 16.82 | 8.00 | 18.22 | 7.78 | 15.41 | 8.00 |  | 9.39E-03 |
| Disability / Insurance | Disability** | 13.59 | 3.06 | 12.92 | 2.87 | 14.26 | 3.10 |  | 1.06E-03 |
|  | Insured Male** | 91.75 | 3.15 | 92.45 | 3.03 | 91.05 | 3.13 |  | 9.59E-04 |
|  | Insured Female** | 93.87 | 2.73 | 94.39 | 2.57 | 93.35 | 2.80 |  | 5.14E-03 |
|  | Insured Black** | 91.42 | 4.24 | 92.16 | 3.03 | 90.69 | 5.10 |  | 1.03E-02 |
|  | Insured Asian | 92.14 | 5.51 | 92.68 | 4.23 | 91.57 | 6.57 |  | 1.45E-01 |
|  | Insured Hispanic | 85.66 | 8.26 | 86.26 | 7.25 | 85.06 | 9.15 |  | 2.83E-01 |
|  | Insured White** | 94.50 | 2.43 | 95.21 | 2.09 | 93.80 | 2.55 |  | 1.33E-05 |
|  | Medicaid** | 22.44 | 7.01 | 20.88 | 6.36 | 24.00 | 7.30 |  | 9.23E-04 |

Full risk factor description is available in Table 1. \*\* denotes statistically significant.

### COVID-19 infection and mortality are higher among African Americans

We have also extracted data on all the states with respect to race distribution (see Table S8). As of April 16, 2020, we have observed that African Americans, as defined in the reports, have a higher rate of COVID-19 infection and a higher death rate (See Table 3). The number of African Americans infected by COVID-19 is 64,605 (1,981 cases per Million) with the number of deaths reaching 6,181 (211 deaths per Million), while the number of Whites, as defined in the reports, is 104,914 (658 cases per Million) infected and 9,806 (76 deaths per Million) dead leading to a disproportional percentage of African Americans infected (p < 0.0001) by COVID-19 and dead (p<0.0001) as the result.

**Table 3.** Rate of COVID-19 positive cases and related deaths with respect to race in the United States as of April 16, 2020.

| **Race** | **Infection Rate (per Million)** | **Death Rate (per Million)** |
| --- | --- | --- |
| **African American** | 1,981 | 211 |
| **White** | 658 | 76 |
| **Latino** | 947 | 82 |
| **Asian** | 390 | 52 |

## DISCUSSION

Our analysis highlights that counties with a higher total population, more diverse demographics, higher education, and income level are at a higher risk of COVID-19 infection; however, counties with a smaller population, higher disability rates, and higher poverty levels have a higher rate of mortality. The conflicting results for counties’ population could be related to the population density, easier access to the high quality of healthcare, and more experience managing the COVID-19 infection due to the higher number of patients. One can argue that counties with fewer residents have a higher rural population. Studies have shown that there are significant differences in the overall healthcare assessment of rural populations as compared to urban populations.^16^ Our observation is also aligned with a recent analysis of health differences in 3,053 US counties, showing that rural areas are more likely to have poorer health outcomes.^17^ The association of poverty and disability makes the conclusion of this study more complex and beyond the analysis of social determinants. By adding the interaction terms in the linear regression model (Death Rate ∼ Poverty + Disability + Poverty:Disability) of Death Rate, we do not observe a significant interaction (p =0.469), suggesting these two variables could be independent in their contribution to the risk of mortality. Populations with a higher disability^18^ and lower median income^19^ might be less mobile, have more comorbidities,^20,21^ and also less likely benefit from timely high-quality care^22^ and high-quality nutrition; all of these factors could be equally important to combating this pandemic.^23,24^ Furthermore, our analysis of the preliminary data on mobility, given the recent social distancing guidelines, corroborate the impact of this intervention on lowering the infection rate and death.

Our findings highlight that race (especially Black) is a risk factor for the infection. To further our understanding of the impact of race, we performed an additional comparative analysis using ANOVA and found that counties with fewer than median non-Hispanic Whites (Group 1: percentage of non-Hispanic Whites ≤39.7%) had a significantly lower total population (p<5.2E-15) than counties with more than median non-Hispanic Whites (Group 2: percentage of non-Hispanic Whites >39.7%); however the rate of mortality is significantly higher (p<0.003) while the rate of infection is significantly lower(p<4.4E-13) in this group; Table S9 includes additional details. Finally, access to insurance was a protective factor in terms of mortality from COVID-19, but access to insurance did not significantly associate with the rate of infection. Comparison of counties with higher or lower than median death rates provided further evidence of the association of lower median income, and higher poverty levels across all the races with mortality. The counties with a higher death rate also had a higher percentage of people on Medicaid. The latter is expected since Medicaid is significantly associated with the rate of poverty (Pearson correlation: 0.769, p<0.005) as well as the rate of disability (Pearson correlation: 0.428, p<0.005). Descriptive statistics of data from all the states also corroborates that African Americans might be disproportionally affected by this pandemic as of April 16, 2020. This observation is consistent with the H1N1 pandemic, where studies have shown evidence of racial and ethnic disparities in the population affected in terms of exposure, severity, and mortality of the disease.^7,8^ Finally historical data have taught us that minorities and people of color tend to be more affected by different diseases.^25–28^

Implications of the results from this study highlight the value of the targeted interventions, as different counties, even within the same state, may have different characteristics and different needs. Furthermore, as the association between COVID-19 related fatality and infection is different among different race and health status, it is important to further study the impact of the immune system and immune-boosting strategies in the at-risk population (such as people with certain disabilities or those residing in elderly community centers), as preventive measure along with other measures based on social distancing guidelines and the ability to work from home.

This is the first systematic study on the racial, health, and economic disparity, as well as education, mobility, and COVID-19 infection in the US with the available data from the most severely impacted states. Our study had several limitations; the data was not granular, and we had missingness, especially for smaller and less populated counties. Access to the infected patient information and mortality data was not possible, and only aggregated data were used. Furthermore, many states claimed difficulties in reporting racial/ethnic demographic data due to patients opting-out of providing their racial identification. The lack of clarity resulted in partially reported data for the death and case rate per million reported in this article, due to some states reporting on racial data for one, two, or all the racial variables specified in this study. The infection rate estimate may be underrepresented, as some individuals may have mild symptoms but lacked clinical validation of the infection. Finally, our in-depth analysis was based on only seven states, leading to conclusions that may not be generalizable to other regions.

## Data Availability

Data sources in this study include, 1) publicly available data from USAfacts and the US Census Bureau for COVID-19 cases and county-level demographic data 2) COVID-19 data reported by each state on their department of health websites, 3) State Population by Race/Ethnicity data, and 4) mobility data extracted from Google. All information on the source (and link) of the data used is provided in the manuscript.

https://usafacts.org/visualizations/coronavirus-covid-19-spread-map/

https://data.census.gov/cedsci/table?q=Computer&tid=ACSST1Y2018.S2801&t=Telephone,%20Computer,%20and%20Internet%20Access

https://www.governing.com/gov-data/census/state-minority-population-data-estimates.html

https://worldpopulationreview.com/states/black-population-by-state/

https://www.google.com/covid19/mobility/

## Funding

This study had no specific funding.

## Notes

**Conflict of Interest:** The authors declare no competing interests.

### Competing Interest Statement

The authors have declared no competing interest.

### Funding Statement

Source of Funding: This study had no specific funding. RZ and VA have funding support from the Geisinger Health Plan Quality Fund as well as the National Institute of Health R56HL116832 (sub-award) during the study period. The funders had no role in study design, data collection, and interpretation, or the decision to submit the work for publication.

